# IgA autoantibodies target pulmonary surfactant in patients with severe COVID-19

**DOI:** 10.1101/2021.02.02.21250940

**Authors:** Tobias Sinnberg, Christa Lichtensteiger, Omar Hasan Ali, Oltin T. Pop, Mara Gilardi, Lorenz Risch, David Bomze, Philipp Kohler, Pietro Vernazza, Werner C. Albrich, Christian R. Kahlert, Silvio D. Brugger, Marie-Therese Abdou, Carl Zinner, Alexandar Tzankov, Martin Röcken, Lukas Kern, Martin H. Brutsche, Hubert Kalbacher, Ana Velic, Boris Maček, Josef M. Penninger, Matthias S. Matter, Lukas Flatz

## Abstract

Complications affecting the lung are hallmarks of severe coronavirus disease 2019 (COVID-19). While there is evidence for autoimmunity in severe COVID-19, the exact mechanisms remain unknown. Here, we established a prospective observational cohort to study lung specific autoantibodies (auto-Abs). Incubation of plasma from severe COVID-19 patients with healthy human lung tissue revealed the presence of IgA antibodies binding to surfactant-producing pneumocytes. Enzyme-linked immunosorbent assays (ELISA) and protein pull-downs using porcine surfactant confirmed the presence of auto-Abs binding to surfactant proteins in severe COVID-19 patients. Mass spectrometry and ELISAs with recombinant proteins identified IgA auto-Abs that target human surfactant proteins B and C. In line with these findings, lungs of deceased COVID-19 patients showed reduced pulmonary surfactant. Our data suggest that IgA-driven autoimmunity against surfactant may result in disease progression of COVID-19.

## INTRODUCTION

The Coronavirus disease 2019 (COVID-19), caused by infection with severe acute respiratory syndrome coronavirus 2 (SARS-CoV-2), has rapidly evolved into a global pandemic with grave socio-economic implications that affects every country worldwide.^1,2^ To date more than 100 million cases of infection with SARS-CoV-2 and over 2 million deaths have been reported across the globe.^3^ While age and comorbidities have been established as predictors for clinical outcome,^4^ there is a lack of validated factors that precede disease progression. Particular susceptibility factors, such as HLA haplotype,^5^ blood type,^6^ and genetic polymorphisms of SARS-CoV-2 target receptors including angiotensin converting enzyme 2 (ACE2)^7^ have been proposed. Yet, they are not utilized in standard clinical management of COVID-19.^8^

Recent publications highlight a potential role of autoimmunity associated with COVID-19 severity, as many patients develop symptoms that resemble autoimmune diseases, such as antiphospholipid syndrome (APS), rheumatoid arthritis, and myositis.^9-11^ Woodruff et al. have shown a surge of B-cell responses that resemble those found in systemic lupus erythematosus (SLE) and further revealed that high concentrations of neutralizing SARS-CoV-2 antibodies are associated with higher mortality.^12^ Bastard et al. reported that autoantibodies (auto-Abs) against the type I-interferons interferons (IFN) α2 and IFN-ω inhibit the immune response against SARS-CoV-2, significantly increasing the likelihood of disease progression.^13^ In line with these findings, an emerging report by Yang et al. demonstrates that severely ill patients develop various auto-Abs against immunomodulatory proteins that potentially perturb antiviral immune responses.^14^ Additionally, several studies have shown significant elevations of APS-associated auto-Abs in severely ill patients, additionally explaining their observed hypercoagulability and vascular inflammation.^11,15^ A further indication for the role of autoimmunity is the significant reduction of mortality in severe and critically ill COVID-19 patients by administration of dexamethasone,^16^ and preventing deterioration of disease using tocilizumab.^17^ Consequently, immunosuppression with dexamethasone has been widely established as standard treatment of severe illness^18^ and other immunosuppressive drugs, such as colchicine and cyclosporine, show benefits in preliminary study data.^19,20^

While there is strong evidence for the role of autoimmunity in severe COVID-19, the exact mechanisms have not been elucidated. Kanduc et al. have demonstrated amino acid sequence similarities of the spike glycoprotein of SARS-CoV-2 and human surfactant-related proteins, suggesting a cross-reactivity of immune responses due to antigen mimicry.^21^ Additional investigations reveal a higher rate of shared protein epitopes between SARS-CoV-2 and the human proteome than with other viruses, further supporting this finding.^22,23^ In a recent paper, Zuniga et al. have shown IgG directed against annexin-2, a phospholipid-binding protein of the lung vasculature, is elevated in severe COVID-19, indicating antibody cross-reactivity.^24^ While autoimmune antibodies with cross- reactivity have been proposed, a lung cell specific target has not yet been shown.

Here, we present that severe COVID-19 is significantly associated with an IgA-driven autoimmune response that targets surfactant proteins in type 2 pneumocytes. This may result in diminished surfactant and, thus, could drive progression of disease severity. We established a prospective observational study and collected plasma samples of COVID-19 patients with varying disease severity. Using immunofluorescence, we observed IgA bound to pulmonary surfactant in plasma of only severely ill COVID-19 patients. We then performed enzyme-linked immunosorbant assays (ELISAs) coated with poractant alfa, a natural surfactant extracted from porcine pulmonary tissue, which revealed significant binding of immunoglobulins (Ig). By mass spectrometry, we were able to identify surfactant proteins as candidate autoantigens. This was confirmed by ELISAs using recombinant human surfactant proteins. Lastly, immunofluorescence of lung tissue of deceased COVID-19 patients showed diminished surfactant. To summarize our findings, we identified IgA auto-Abs targeting surfactant proteins which demonstrate IgA-driven autoimmunity, likely contribuing to compromised blood oxygenation. These data reveal an urgent need to explore the preservation of surfactant proteins by immunosuppression and surfactant replacement in patients with severe COVID-19.

## RESULTS

### Damaged lung tissue in COVID-19 shows autoimmune gene expression signatures

Most COVID-19 patients with fatal outcome develop diffuse alveolar damage (DAD), a life-threating destructive lung disease that leads to disturbed pulmonary function with reduced gas exchange.^25^ Several pathologies, such as pulmonary edema, microthrombosis and fibrosis lead to respiratory failure of patients with DAD. DAD from COVID-19 patients (COVID-19 DAD) show less pronounced exudative changes and hemorrhages compared to non-COVID-19 DAD, as well as more microthromboses, pulmonary thromboembolisms and new vessel growth (intussusceptive angiogenesis), indicating additional drivers of pathology.^25-27^ In addition, COVID-19 is characterized by an exaggerated immune response in the lung.^28^ Thus, we hypothesized that the respiratory failure of COVID-19 patients may be further aggravated by auto-Abs against self-antigens evoked by an exaggerated immune activation. To understand if molecular pathways involved in autoimmunity are present in lung tissue of severe COVID-19 patients (n=11), we performed gene expression analysis with a panel specifically designed to analyze autoimmune disease (HTG EdgeSeq Immune Response Panel). As a control group, we used patients with a DAD (n=10) caused by other reasons such as drug-toxicity, lung surgery or myocardial infarction (**Fig. 1a**; **Supplementary Table 1**). Unsupervised hierarchical clustering by *k*-means revealed clear grouping of COVID-19 patients and non-COVID-19 patients in spite of both cohorts presenting with DAD (**Fig. 1b**). To explore, if networks of autoimmune diseases are activated in COVID-19 patients, we performed gene set enrichment analyses. To that end, we searched the Kyoto Encyclopedia of Genes and Genomes database (KEGG; https://www.genome.jp/kegg/pathway.html) for autoimmune diseases and selected “systemic lupus erythematosus (SLE)” and “autoimmune thyroid disease (ATD)”. To date, these autoimmune diseases are the only ones in KEGG that entail a substantial production of auto-Abs. While both pathways showed significant expression enrichment in lung tissue of COVID-19 autopsy samples (**Fig. 1c**), only the enrichment compared to SLE remained significant after false discovery rate (fdr)-adjustment (SLE normalized enrichment score: 1.6, *P*=0.0008, fdr-adjusted *P*=0.02; vs. ATD normalized enrichment score: 1.9, *P*=0.01, fdr-adjusted *P*=0.10). In conclusion, these data show that networks of autoimmune disorders with the production of auto-Abs are enriched in COVID-19 patients.

**Fig. 1:**
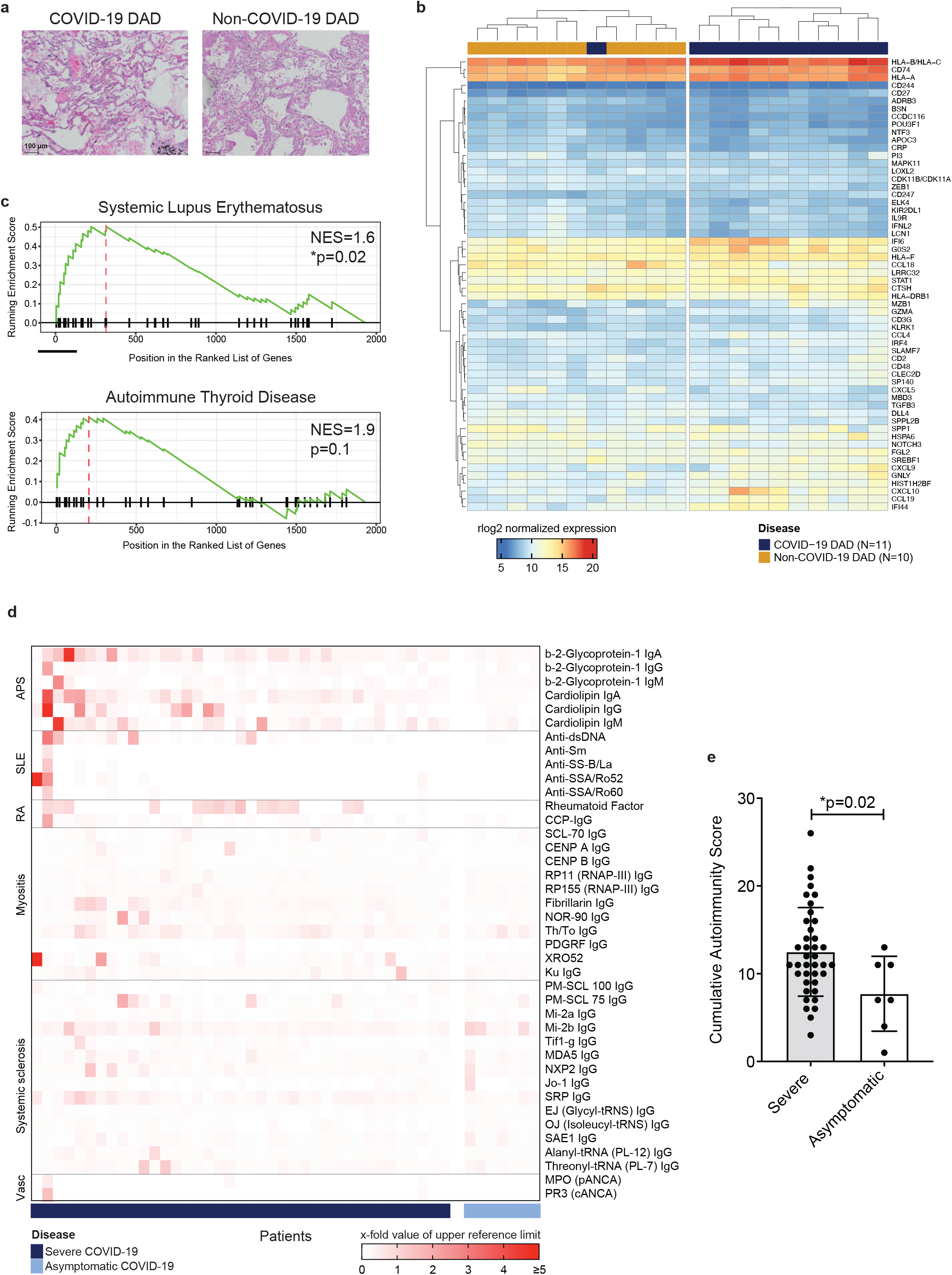
COVID-19 patients with severe illness have a distinct mRNA expression pattern in damaged alveolar tissue and elevated autoantibodies in their blood. **a**, Representative images of diffuse alveolar disease (DAD) from COVID-19 patients and non- COVID-19 patients show hyaline membrane formation, desquamation and beginning of septal fibrosis. Hematoxylin and eosin staining. **b**, RNA sequencing data of DAD resulting from COVID-19 (n=11) and other causes (n=10) reveals a clustered upregulation of genes. **c**, Gene set enrichment analysis shows a significant correlation of the systemic lupus erythematosus enrichment pattern and COVID-19 (*P*=0.02). A trend can be observed for autoimmune thyroid disease. **d**, Heatmap (center panel) displaying fold-levels of autoantibodies (auto-Abs) relative to the upper reference limits of the diagnostic laboratory assays, in descending order (left to right). The left panel shows autoimmune diseases, while the right panel lists associated auto-Abs. The bottom panel indicates the individual patients, with severe (marine blue, n=39) or asymptomatic (light blue, n=7) COVID-19. Abbreviations: APS = antiphospholipid syndrome, RA = rheumatoid arthritis, SLE = systemic lupus erythematosus, Vasc = vasculitis. **e**, Comparison of the cumulative autoimmunity scores derived from the median value of each antibody. Patients with severe COVID-19 score (n=39) significantly higher than asymptomatic patients (n=7, *P*=0.02). Score calculation is described in the Methods section.

### Clinical study design

In order to identify auto-Abs in patients with COVID-19 we enrolled N=46 individuals in a prospective monocentric observational study and collected plasma samples. The median age was 61 years (interquartile range 55-70 years), and 32 (70%) were male. Patient characteristics are provided in **Supplementary Table 2**. Patients were divided into subgroups by COVID-19 severity based on clinical criteria.^29^ In summary, we included 26 (57%) patients with critical, 13 (28%) with severe, and 7 (15%) with asymptomatic illness. For simplicity, patients with critical and severe disease will be referred together as severe COVID-19. Additionally, we included plasma samples from healthy, non- infected patients for control measurements. Details on patient recruitment are provided in the Methods section.

### Autoimmune antibodies are elevated in severe COVID-19

We assessed the presence of autoimmune-disease specific auto-Abs in COVID-19 patients and found several auto-Abs that were elevated in individuals suffering from severe COVID-19 compared to asymptomatic patients (**Fig. 1d**). Using the median antibody values of all cohorts, we calculated a cumulative autoimmunity score (CAS). As expected, the CAS in severe COVID-19 patients was significantly higher than in asymptomatic patients (*P*=0.02, two-tailed t-test, **Figure 1e**). In line with the literature,^14^ our data showed elevated auto-Abs in severe COVID-19 compared to asymptomatic patients, suggesting a vigorous auto-Abs response associated with disease severity.

### Severe COVID-19 patients develop autoantibodies against proteins in type II pneumocytes

Given that we have detected autoimmune gene expression signatures, as well as auto-Abs, we next determined the reactivity of auto-Abs in plasma samples from severe COVID-19 patients to any lung protein using indirect immunofluorescence. Here, we inoculated plasma of severe COVID-19 patients and healthy control patients with normal human alveolar lung tissue. We discovered specific binding of IgA antibodies to distinguished areas of lung tissue (**Fig. 2a**). IgG were discoverable to a lesser extent (**Supplementary Fig. 1**). Given that type II pneumocytes are the primary pulmonary target of SARS-CoV-2,^30^ we assessed mRNA expression from various lung alveolar cells using previously published data.^31^ This revealed a characteristically high gene expression of surfactant proteins C (SFTPC), B (SFTPB), A1 and A2 in type II pneumocytes (**Fig. 2b**). To examine, whether COVID-19 patients showed IgA and IgG derived autoreactivity against surfactant-producing cells, we performed immunofluorescence and co-stained for surfactant protein A as a surrogate marker for type II pneumocytes. This showed strong co-localization of IgA and surfactant (**Fig. 2c**), and, to a lesser extent, of IgG and surfactant (**Supplementary Fig. 2**). Thus, our data indicate that predominantly IgA auto-Abs bind to surfactant proteins in severe COVID-19 patients.

**Fig. 2:**
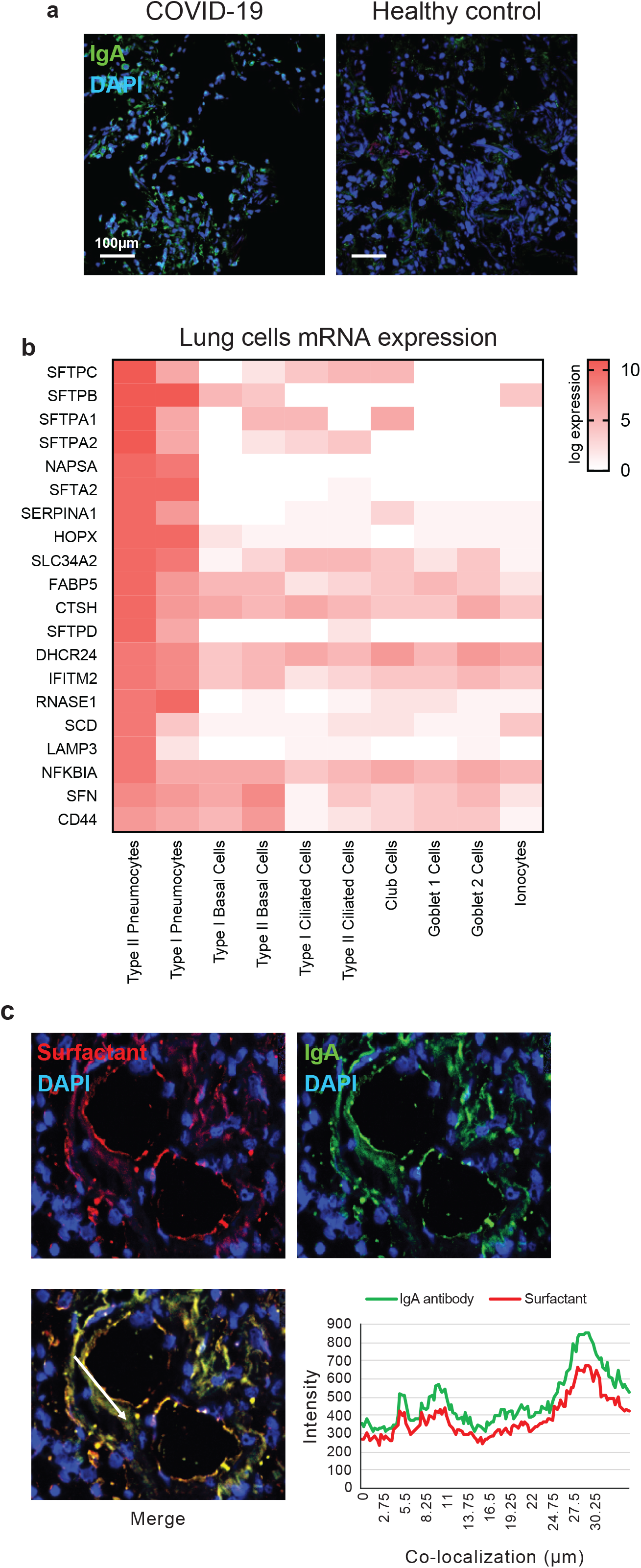
Immunoglobulins of severe COVID-19 patients co-localize with surfactant in lung tissue. **a**, Indirect Immunofluorescence of lung tissue shows that IgA antibodies are present in diffuse alveolar damage of COVID-19 patients. **b**, RNA-seq expression of type II pneumocytes, the main producer of surfactant, shows abundant overexpression of surface proteins C (SFTPC), B (SFTPB), A1 (SFTPA1) and A2 (SFTPA2). **c**, Immunofluorescence staining of lung tissue with serum from severely ill COVID-19 patients shows co-localization of IgA binding with surfactant. Co-localization of IgA and the target protein is visualized in the right lower graph.

### IgA autoantibodies target surfactant protein B and C in patients with severe COVID-19

In order to identify the target surfactant protein, we performed ELISA coated with poractant alfa. Poractant alfa is an extract of porcine lung surfactant that consists of various phospholipids and phospholipid-binding proteins, and is used as an intratracheal rescue therapy for infants with acute respiratory lung distress syndrome (ARDS).^32^ ELISA results showed a significant presence of IgA (*P*=0.006, Mann-Whitney U test, **Fig. 3a**) and IgG (*P*=0.001, **Supplementary Fig. 3**) directed against poractant alfa in the plasma of severe COVID-19 patients compared to asymptomatic disease. Next, we identified protein components of poractant alfa using mass spectrometry, which confirmed the presence of SFTPB and SFTPC (**Fig. 3b**). To identify putative antigens, we prepared pull-down affinity columns: here we used pooled and immobilized Ig from severe COVID-19 patients (n=14) and uninfected healthy controls (n=5), after their incubation with poractant alfa. The extract was analyzed by mass spectrometry to determine the Ig-captured proteins. Interestingly, SFTPB was the most abundant protein with high enrichment (≥ 10-fold) for captured proteins from plasma of severe COVID-19 patients (**Fig. 3c**). Next, we used recombinant proteins of SFTPB and SFTPC to determine the presence of autoreactive IgA and IgG in severe COVID-19 patients. The results showed a highly significant difference of IgA against SFTPB (severe vs. asymptomatic, *P*=0.001, Mann-Whitney U test) and for SFTPC (severe vs. asymptomatic, *P*=0.004) (**Fig. 3d**). Conversely, they showed no significant difference for autoreactive IgG (**Supplementary Fig. 4**).

**Fig. 3:**
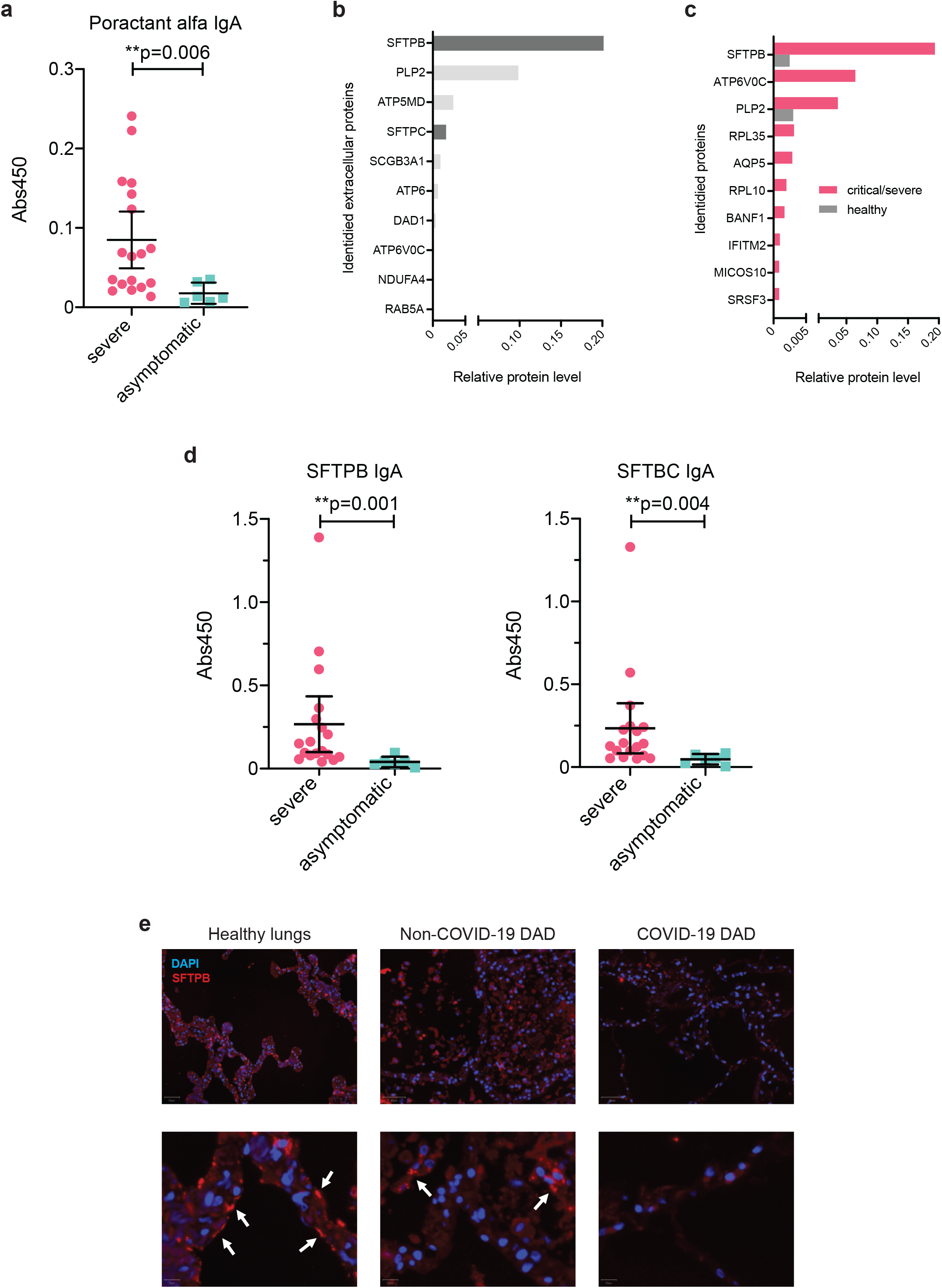
Severe COVID-19 is associated with elevated IgA against surfactant proteins B and C, and shows diminished alveolar surfactant. **a**, ELISA with poractant alfa (Curosurf®) demonstrates that significantly more IgA is binding in plasma from severe COVID-19 (n=18) compared to asymptomatic patients (n=6; mean with 95% CI, *P=*0.006). **b**, Mass spectrometry of poractant alfa lists abundance of surfactant binding proteins B (SFTPB) and C (SFTPC). **c**, Pull-downs of poractant alfa reveal immunoglobulins binding to SFTPB predominantly in severe COVID-19 samples. **d**, ELISA with recombinant proteins show that patients with severe COVID-19 (n=18) have significantly more IgA against SFTPB (*P*=0.001) and significantly more IgA against SFTPC (*P*=0.004) than asymptomatic patients (n=6) in the blood (mean with 95% CI). **e**, Indirect immunofluorescence with healthy lung tissue indicates reduced amounts of surfactant in non-COVID-19 diffuse alveolar damage (DAD) than in COVID-19 DAD.

### Surfactant is diminished in lungs of deceased COVID-19 patients

SARS-CoV-2 primarily infects cells via their angiotensin converting enzyme 2 (ACE2) receptors.^33^ By infecting alveolar type II epithelial cells, which are rich in ACE2 receptors on their surface, SARS-CoV-2 directly influences the function and production of surfactant proteins.^30^ Our data of auto-Abs against surfactant protein suggest an additional mechanism of surfactant disturbance. To investigate, whether surfactant in lung tissue of COVID-19 patients is affected, we assessed its presence in deceased COVID-19 patients with DAD and compared it to patients with non-COVID-19 DAD, and healthy controls. As suggested by previously reported gene expression analysis,^34^ the total amount of surfactant was reduced in COVID-19 patients in comparison to non-COVID-DAD as well as to healthy controls (**Fig. 3e**).

In summary, we identified novel IgA-driven autoimmune response against SFTPB and SFTPC in severe COVID-19 patients, which is associated with diminished surfactant. Our data suggest that a vigorous B-cell response against self-antigens can corrupt crucial components for alveolar gas exchange, possibly explaining progression of COVID-19 to ARDS. We suggest further mechanistic exploration of this hypothesis.

## METHODS

### Gene expression analysis

Samples were included based on quality control criteria by the manufacturer (HTG Molecular Diagnostics, Tucson, AZ). These require that the percentage of overall reads allocated to the positive process control probe per sample is less than 28%, the read depth is at least 750000 and the relative standard deviation of reads allocated to each probe within a sample is greater than 0.094. In addition, only samples with a known postmortem interval were preserved for differential analysis. Differential expression analysis was conducted in R version 4.0.3 (R Project for Statistical Computing, Vienna, Austria) with the DESeq2 package using default settings. Count estimates were normalized with the median ratio method. Differential gene expressions for the contrast of COVID-19 patients with DAD vs. control patients with other DAD were modelled with a negative binomial distribution and subjected to a Wald significance test. Prior to heatmap visualization, the normalized counts were further $log_2$ transformed using a robust variance stabilization. The heatmap of genes with a | log fold change | > 1 was produced with the complexHeatmap package. Column sample clusters were obtained by k-means clustering with k = 2 and row gene clusters by hierarchical clustering with complete linkage. Functional analysis was quantified via gene set enrichment. All genes in the autoimmune panel were pre-ranked using the Wald test statistic and submitted to the entire KEGG database of human pathways using the clusterProfiler package.

### Clinical study: patient recruitment, data and blood sample collection

Patient data and blood collection was approved by the Ethics committee of Eastern Switzerland (study ID 2020-01006), and tissue collection by Ethics committee of Northern and Central Switzerland (study ID 2020-00969). The studies were performed in accordance with the Declaration of Helsinki guidelines.^35^ Collection of patient data and samples (serum or plasma) was conducted from April 9, 2020, to May 1, 2020 and tissue collection from March 13 2020 to May 4, 2020. SARS-CoV-2 infection of the severe COVID-19 cohort was confirmed by real-time reverse transcriptase- polymerase chain reaction^36^ of nasopharyngeal swab samples. Infection of asymptomatic COVID-19 was confirmed with serology of SARS-CoV-2 antibodies, using two independent antibody tests: SGIT flex Covid 19, a lateral flow immunochromatographic assay (LFIA) (Sugentech, Daejeon, South Korea), and Elecsys Anti-SARS-CoV-2, an electro-chemiluminescence immunoassay (ECLIA) (Roche International Diagnostics AG, Rotkreuz, Switzerland). We defined a positive result as elevated SARS-CoV-2 IgG in the LFIA in the acute phase with confirmation by IgG in the ECLIA after 3-4 weeks. All blood samples of severe COVID-19 patients were obtained within the first two weeks of symptom onset. Due to the unknown primary infection of patients with asymptomatic COVID-19, blood draw at an unknown timepoint is assumed.

### Plasma isolation from whole blood

Plasma samples were isolated from sodium-heparin whole blood (BD Vacutainer® CPT™ tubes, Becton Dickinson, NJ). Briefly, the tubes were centrifuged at room temperature (RT) at 1650 g for 20 minutes at room temperature. The undiluted plasma was then aliquoted and stored at −80 °C for subsequent analysis.

### Antibody analysis

Myositis and systemic sclerosis antibodies (i.e. Mi-2a, Mi-2b, TIF1g, MDA5, NXP2, SAE1, Ku, PM- SCL100, PM-SCL75, Jo-1, SRP, PL-7, PL-12, EJ, OJ, Ro5 for myositis; Scl-70, CENPA, CENPB, RP11 (RNAP-III), RP155 (RNAP-III), Fibrillarin, NOR-90, Th/To, PM-Scl100, PM-Scl75, Ku, PDGRF, Ro-52 for systemic sclerosis) were determined via immunoblot using the EuroBlotOne test system (Euroimmun AG, Lübeck, Germany), respectively, following the manufacturer’s instructions. Autoantibodies against Cardiolipin, β-2-glycoprotein 1, double stranded DNA (dsDNA), cyclic citrullinated peptide (CCP), SSA1, SSA2, SSB, Sm, PR3, and MPO were determined by fluorescence enzyme immunoassay on a Unicap 250 analyzer (Thermo Fisher Scientific, Waltham, U.S.A.). Rheumatoid factor (RF) was determined by turbidimetry (COBAS 6000, Roche Diagnostics, Rotkreuz, Switzerland). Measurements were performed at the Labormedizinisches Zentrum Dr. Risch (Buchs, St. Gallen, Switzerland), an ISO 17025:2018 accredited medical laboratory. The cumulative autoimmunity score (CAE) for each patient was generated in two steps: First, we calculated the median value of all the analyses results of a given autoantibody. Second, we determined for each patient, if the result is above (score of “1”) or below (score of “0”) that value. The CAE represents the sum of those scores across all autoantibodies for any given patient.

### Indirect immunofluorescence

Four serial frozen sections from 4 lung tissues from patients without COVID-19 infection were washed two times with PBS 0.1% Tween 20, for 5 min each. In a first step one section from each lung tissue was then incubated for 30 minutes at RT with plasma from severe COVID-19 patients (plasma pooled from three patients and diluted 1:3 in PBS), and another with serum from healthy donors (pooled from three individuals, diluted 1:3 in PBS), and two slides with PBS only (negative controls). After washing, one of the of the control slides was then incubated with sterile PBS for 30 min at room temperature, while the remaining three slides were incubated with anti-IgA FITC (Dako, catalog no. F0204; dilution 1:50) or anti-IgG FITC (Dako, catalog no. F0202; dilution 1:50) for 30 minutes at RT. Slides were then counterstained with DAPI (ThermoFisher, catalog no. D1306) and mounted using fluorescence mounting medium (Dako, catalog no. S3023). Slides were imaged and assessed using a Zeiss LSM 710 laser scanning microscope (Carl Zeiss AG, Oberkochen, Germany).

### Co-localization immunofluorescence staining

Fresh frozen lung tissues were incubated with the above described patients’ plasma pools diluted 1:3 for 30 min RT. Immunofluorescence staining was performed as previously described.^37-39^ Briefly, anti-surfactant (diluted 1:100 ab51891, Abcam), anti-IgA FITC and anti-IgG FITC primary antibodies (diluted 1:50, respectively F0316-F0202 Dako) were used. Then, the incubation with the serum secondary antibody alexa-fluor 647 (Invitrogen) was done. DAPI incubation for 10 minutes has been performed to stain the nuclei. Samples were mounted in prolong gold anti-fade mounting medium (Invitrogen) and scanned by confocal microscope (Ti2, Nikon, Tokyo, Japan). Images were analyzed using QuPath software^40^ as previously reported.^37^ Intensity fluorescence profile has been analyzed by Nikon software to determine the co-localization of the signals as previously reported.^38^

### SFTPB immunofluorescence staining

We compared the presence of SFTPB in formalin-fixed paraffin-embedded lung tissues obtained from deceased patients with COVID-19 DAD, non-COVID-19 DAD and normal lungs as controls. Tissues were processed as previously described for immunofluorescence staining.^37-39^ Briefly, antigen retrieval was performed, and tissues incubated with PBS 0.1% Tween-20. Staining using anti-SFTPB antibodies diluted 1:100 (NCL-SPPB) was done in blocking buffer for 1h at RT. Alexa-fluor 647 (Invitrogen) was applied as secondary antibody. Controls for secondary antibody specificity were generated by substituting the primary antibody with blocking buffer only. All samples were incubated with DAPI for 10 minutes and mounted in prolong gold anti-fade mounting medium (Invitrogen) and scanned with the Ti2 confocal microscope (Nikon, Tokyo, Japan).

### Single cell RNA data analysis

Single-cell RNA sequencing data of healthy lungs were obtained from the LungMap project^31^ via the ToppCell portal (https://toppcell.cchmc.org/). Counts were normalized to log2-transcripts per million (TPM) and compared across the 7 major epithelial cell types: type-1, and type-2 alveolar-, basal-, ciliated-, club-, and goblet cells, as well as ionocytes. Housekeeping genes were excluded from the analysis. Cell-specific genes were defined as those with overall expression log2-TPM > 5 and at least 4-fold higher expression in one or two particular cell types compared to all other cell types.

### Immunoglobulin isolation from COVID-19 plasma samples

Plasma samples from 14 critical/severe COVID-19 patients and 5 asymptomatic patients were pooled and immunoglobulins were isolated from 1 ml each of the plasma pools using 4 mL protein L coupled agarose resin (Capto™ L, Cytiva Life Sciences, Amersham, UK) packed into 15 mL empty polypropylene columns (Chromabond, Machery-Nagel, Düren, Germany). After extensive washing with PBS-T (0.05% Tween 20) the immunoglobulins were eluted in 0.1 M glycine buffer pH 2.5. 50 % of the isolated immunoglobulins from the pools were individually and chemically cross-linked onto 1 g of CNBr-activated sepharose (CNBr-activated Sepharose® 4B, Cytiva) as recommended by the manufacturer and packed into 15 mL empty polypropylene columns (Chromabond, Machery-Nagel). The protein content of 0.33 mL Curosurf® (poractant alfa, Chiesi Farmaceutici, Parma, Italy) was immuno-purified after acetone precipitation in 15 mL of PBS-T per column by overnight binding at 4°C using a peristaltic pump (ISM597A, Ismatec, Wertheim, Germany) and a circuit flow rate of 0.5 mL/min, washing with 100 ml of PBS-T and elution in glycine buffer pH 2.5.

### Protein identification by mass spectrometry

A SDS PAGE short gel purification was run with either acetone-precipitated 0.2 mL Curosurf® or the immuno-purified and eluted proteins and in-gel digestion with Trypsin was conducted as described previously.^41^ Extracted peptides were desalted using C18 StageTips^42^ and subjected to LC-MS/MS analysis. LC-MS/MS analyses were performed on an Easy-nLC 1200 UHPLC (Thermo Fisher Scientific, Waltham, MA) coupled to an QExactive HF Orbitrap mass spectrometer (Thermo Fisher Scientific) as described elsewhere,^43^ or an Orbitrap Exploris 480 mass spectrometer (Thermo Fisher Scientific) as previously described.^44^ Peptides were eluted with a 60 min segmented gradient at a flow rate of 200nl/min, selecting 20 most intensive peaks for fragmentation with HCD. The MS data was processed with MaxQuant software suite v.1.6.7.0^45^ to measure the iBAQ which was used to calculate the relative protein level (riBAQ=iBAQ/(ΣiBAQ)). Database search was provided against pig (121817 entries) and human (96817 entries) UniProt database using the Andromeda search engine.^46^

## Supporting information

STROBE Checklist

## Data Availability

The data that support the findings of this study are available from the corresponding author, L.F., upon reasonable request.

## ACKNOWLEDGEMENTS

We thank Dorothea Hillmann (Labormedizinisches Zentrum Dr. Risch) for her contributions to laboratory analyses.

## Funding

This research was supported by grants from the Swiss National Science Foundation (PP00P3_157448 to L.F., P400PM_194473 to O.H.A., and 320030_189275 to M.S.M.), the Research Fund of the Kantonsspital St. Gallen (20/20 to L.F.), the Promedica Foundation (1449/M to S.D.B.), and the Botnar Research Centre for Child Health Emergency Response to COVID-19 Grant (to C.Z., M.S.M., and A.T.).

## Competing interests

J.M.P. is founder and shareholder of Apeiron (Vienna, Austria), developing soluble ACE2 as a COVID-19 therapy. J.M.P. has no direct competing interest relating to the paper or data presented in the paper. M.S.M. has received speaker honoraria from Thermo Fisher Scientific (Waltham, MA) and honoraria as an advisory board member from Novartis AG (Basel, Switzerland). All other authors have no competing interests to declare.

**Supplementary information** is available for this paper.

## Notes

### Author Declarations

The study was approved by the Ethics committee of Eastern Switzerland (study ID 2020-01006) and Ethics committee of Northern and Central Switzerland (study ID 2020-00969).

### Summary of Updates

Minor amendment to author names, affiliations and acknowledgements.

