## Supplementary Table 1 for "IgA autoantibodies target pulmonary surfactant in patients with severe COVID-19"

**Supplementary Table 1:** Characteristics of lung tissue samples

| Patient no. | Cause of DAD |
|---|---|
| 1 | COVID-19 |
| 2 | COVID-19 |
| 3 | COVID-19 |
| 4 | COVID-19 |
| 5 | COVID-19 |
| 6 | COVID-19 |
| 7 | COVID-19 |
| 8 | COVID-19 |
| 9 | COVID-19 |
| 10 | COVID-19 |
| 11 | COVID-19 |
| 12 | Post-surgery of lung, lung cancer |
| 13 | Drug toxicity (leukemia, complete remission) |
| 14 | Drug toxicity (leukemia, complete remission) |
| 15 | ICU stay (sepsis, drug toxicity) |
| 16 | Resolved pneumonia |
| 17 | Post myocardial infarction, advanced cancer |
| 18 | Drug toxicity (myelodysplastic syndrome) |
| 19 | Drug toxicity (myelodysplastic syndrome, complete remission) |
| 20 | Drug toxicity or viral infection |
| 21 | Post myocardial infarction, advanced cancer |

Abbreviations: COVID-19 = Coronavirus Disease 2019, DAD = diffuse alveolar damage, ICU = intensive care unit, no. = number
