## Supplementary Table 2 for "IgA autoantibodies target pulmonary surfactant in patients with severe COVID-19"

**Supplementary Table 2:** Patient demographics

| Characteristic | Critical Illness | Severe Illness | Asymptomatic | Total |
|---|---|---|---|---|
| Patient count n (% of total) | 26 (57) | 13 (28) | 7 (15) | 46 (100) |
| Mean Age (y, IQR) | 64 (57-70) | 67 (65-74) | 35 (34-38) | 61 (55-70) |
| Male sex - no. (%) | 20 (77) | 9 (69) | 3 (43) | 32 (70) |
| Respiratory support (%) | 26 (100) | 13 (100) | 0 (0) | 39 (85) |
| Supplementary oxygen only (%) | 1 (4) | 7 (54) | 0 (0) | 8 (17) |
| High-Flow (%) | 0 (0) | 6 (46) | 0 (0) | 6 (13) |
| Non-invasive ventilation (%) | 7 (27) | 0 (0) | 0 (0) | 7 (15) |
| Intubation (%) | 13 (50) | 0 (0) | 0 (0) | 13 (28) |
| ECMO (%) | 5 (19) | 0 (0) | 0 (0) | 5 (11) |
| Arterial Hypertension (%) | 11 (42) | 6 (46) | 0 (0) | 17 (37) |
| Coronary artery disease (%) | 4 (15) | 2 (15) | 0 (0) | 6 (13) |
| Diabetes mellitus Type 2 (%) | 12 (46) | 6 (46) | 0 (0) | 18 (39) |
| Autoimmune disease[1] (%) | 6 (23) | 2 (15) | 0 (0) | 8 (17) |

[1]Autoimmune disease: Graves' disease (n=2), polymyalgia rheumatica (n=1). Abbreviations: ECMO = extracorporeal membrane oxygenation, IQR = interquartile range, y = years.
