## Supplementary Fig. 1 for "IgA autoantibodies target pulmonary surfactant in patients with severe COVID-19"

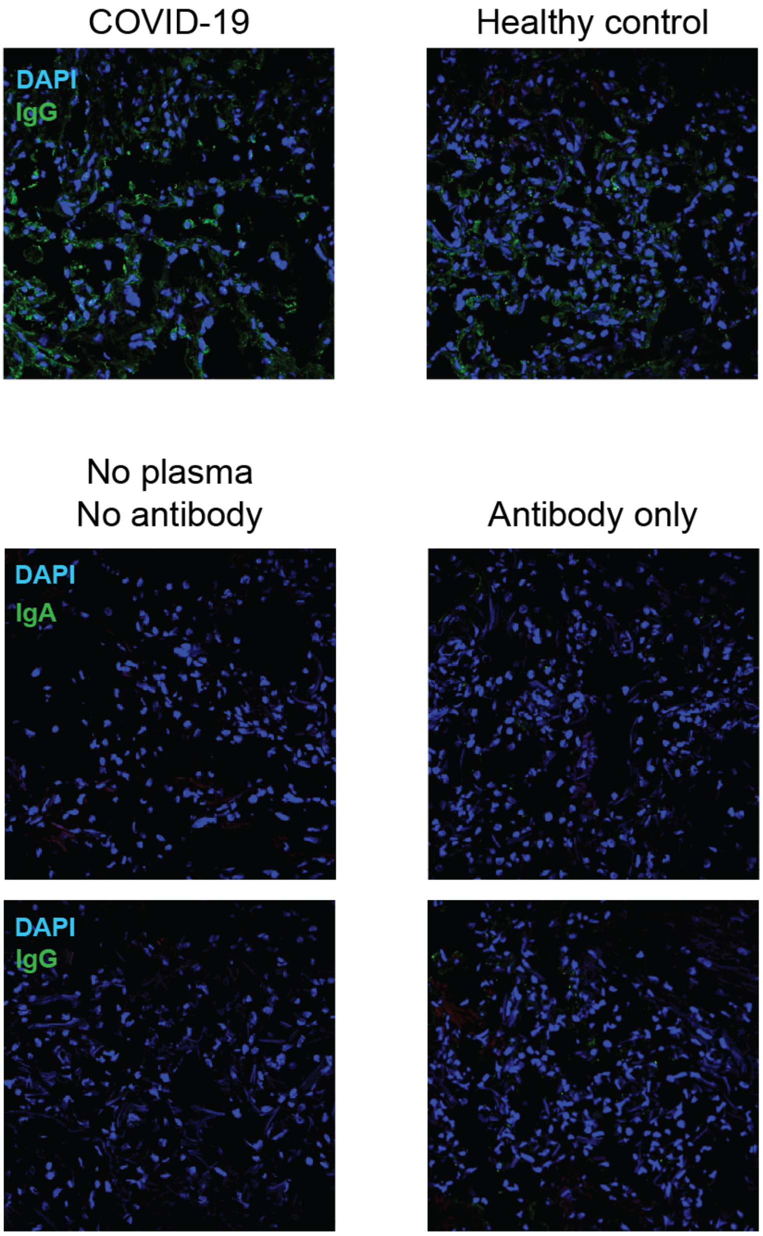


Upper panel: Indirect immunofluorescence of lung tissue shows less binding of IgG than IgA (see **Fig. 2a**). Lower panel: Control antibody stainings of IgA (top) and IgG (bottom).
