## Supplementary Fig. 2 for "IgA autoantibodies target pulmonary surfactant in patients with severe COVID-19"

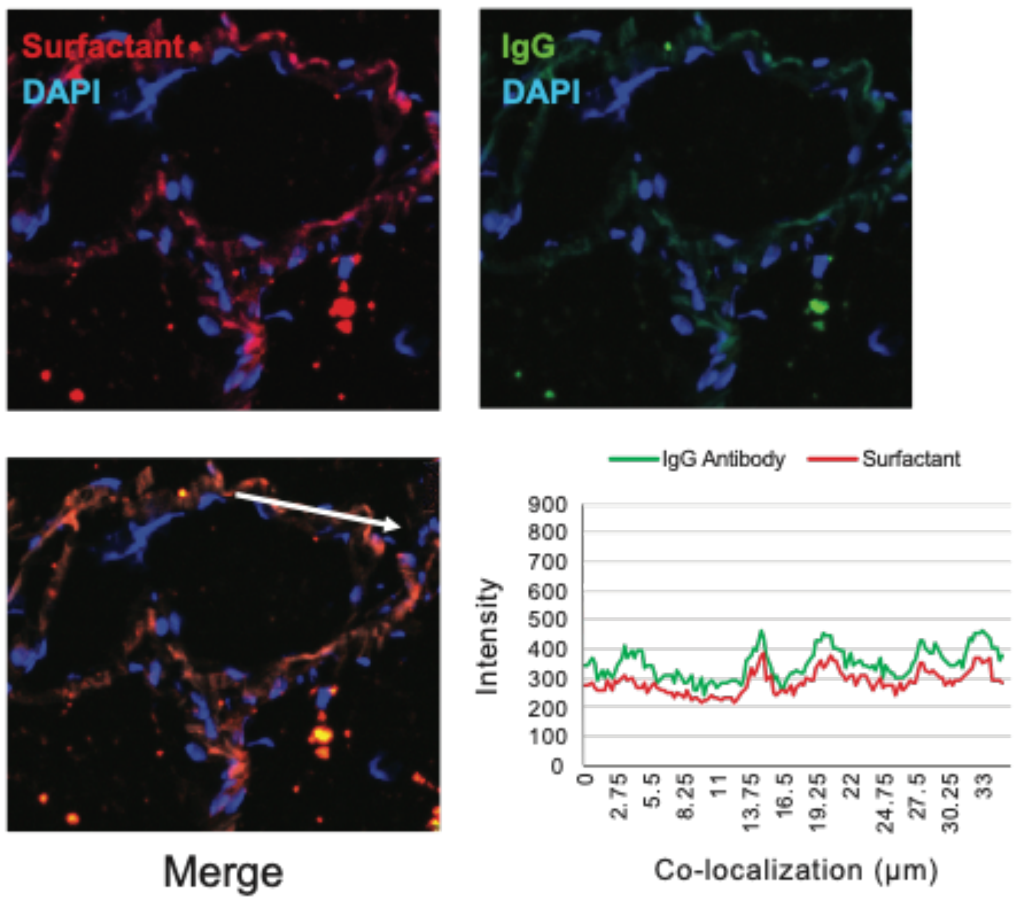


Immunofluorescence depicts colocalization of IgG and surfactant using plasma of severe COVID-19 patients, however less than IgA and surfactant (compare **Fig. 2c**). Colocalization distribution is presented in the lower right graph.
