## Supplementary Fig. 3 for "IgA autoantibodies target pulmonary surfactant in patients with severe COVID-19"

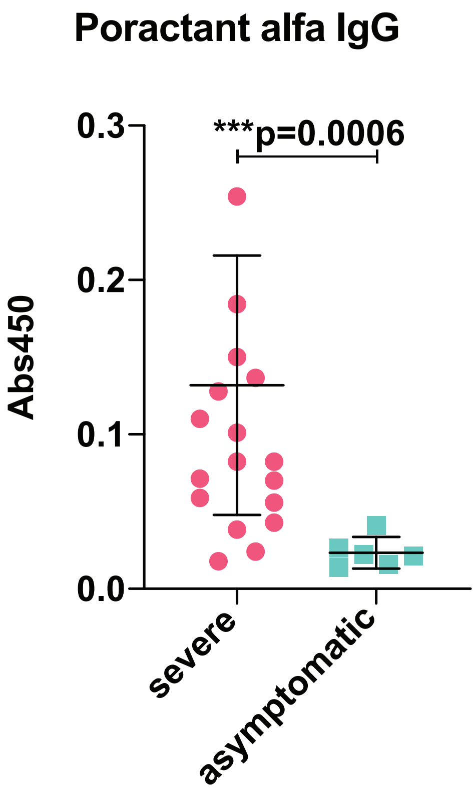


ELISA of IgG against poractant alfa (Curosurf®) shows a significant difference when comparing severe and asymptomatic patients (*P*=0.0006, Mann-Whitney U test).
