## Supplementary Fig. 4 for "IgA autoantibodies target pulmonary surfactant in patients with severe COVID-19"

**Supplementary Figure 4**

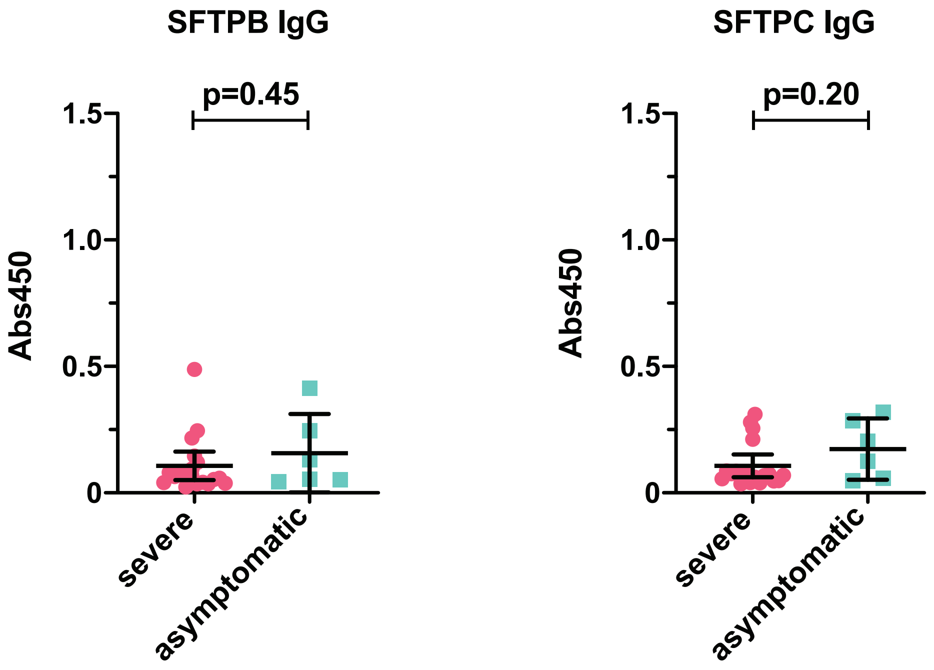


ELISA of IgG against surfactant proteins B (SFTPB) and C (SFTPC) show no significant differences when comparing severe and asymptomatic COVID-19 patients.
